# Safety Outcomes of Mechanical Thrombectomy Versus Combined Thrombectomy and Intravenous Thrombolysis in Tandem Lesions

**DOI:** 10.1101/2023.02.20.23286212

**Authors:** Aaron Rodriguez-Calienes, Milagros Galecio-Castillo, Mudassir Farooqui, Ameer E. Hassan, Mouhammad A. Jumaa, Afshin A. Divani, Marc Ribo, Michael Abraham, Nils H. Petersen, Johanna Fifi, Waldo R. Guerrero, Amer M. Malik, James E. Siegler, Thanh N. Nguyen, Albert J. Yoo, Guillermo Linares, Nazli Janjua, Darko Quispe-Orozco, Wondwossen G. Tekle, Hisham Alhajala, Asad Ikram, Federica Rizzo, Abid Qureshi, Liza Begunova, Stavros Matsouka, Nicholas Vigilante, Sergio Salazar-Marioni, Mohamad Abdalkader, Weston Gordon, Jazba Soomro, Charoskon Turabova, Juan Vivanco-Suarez, Maxim Mokin, Dileep R. Yavagal, Tudor Jovin, Sunil Sheth, Santiago Ortega-Gutierrez

## Abstract

**Background and Purpose:** We aimed to describe the safety and efficacy of mechanical thrombectomy (MT) with or without intravenous thrombolysis (IVT) for patients with tandem lesions (TLs) and whether using intraprocedural antiplatelet therapy (APT) influences MT’s safety with IVT treatment.

**Methods:** This is a sub-analysis of a pooled, international multicenter cohort of patients with acute anterior circulation TLs treated with MT. Primary outcomes included symptomatic intracranial hemorrhage (sICH) and parenchymal hematoma type 2 (PH2). Additional outcomes included hemorrhagic transformation (HT), successful reperfusion (modified Thrombolysis in Cerebral Infarction [mTICI] 2b-3), complete reperfusion (mTICI 3), favorable functional outcome (90-day modified Rankin score [mRS] 0-2), excellent functional outcome (90-day mRS 0-1), in-hospital mortality, and 90-days mortality.

**Results:** Of 691 patients, 599 were included (255 underwent IVT+MT and 344 MT alone). There was no difference in the risk of sICH (aOR=1.43; 95%CI:0.72–2.87; *p=*0.308), PH2 (aOR=1.14; 95%CI:0.57– 2.28; *p*=0.705), and HT (aOR=0.92; 95%CI:0.54–1.57;*p*=0.751) between the IVT+MT and MT alone groups after adjusting for confounders. There was an IVT-by-intraprocedural APT interaction for sICH (*p* _interaction_=0.031). Administration of IVT was associated with an increased risk of sICH in patients who received IV-APT (aOR=3.58; 95%CI:1.17–10.89;*p=*0.025). The IVT+MT group had higher odds of 90-days mRS 0-2 (aOR=1.76; 95%CI:1.05–2.94;*p=*0.030). The odds of successful reperfusion, complete reperfusion, 90-days mRS 0-1, in-hospital mortality, or 90-days mortality did not differ between the IVT+MT vs. MT alone groups.

**Conclusion:** Our study showed that the combination of IVT with MT for TL did not increase the overall risk of sICH, PH2, or overall HT independently of the cervical revascularization technique used. However, intraprocedural IV-ATP during acute stent implantation might be associated with an increased risk of sICH in patients who received IVT prior to MT. Importantly, IVT+MT treatment was associated with a higher rate of favorable functional outcome at 90 days.

## INTRODUCTION

Tandem lesions (TLs) account for about 15% to 30% of large vessel occlusion (LVO) strokes.^1-3^ Mechanical thrombectomy (MT) has demonstrated a safe and effective profile for treating TLs. However, the optimal acute cervical endovascular approaches to optimize outcomes, including angioplasty alone versus angioplasty and stenting, are currently under investigation.^2-7^ Furthermore, the role of intravenous thrombolysis (IVT) in combination with different treatment options is incompletely defined.

In an international survey performed by our group for treating TLs, the use of IVT+MT was controversial.^8^ First, antiplatelet therapy (APT) during endovascular treatment for TL may be safe and associated with lower mortality.^9^ However, when co-administered with IVT in patients undergoing acute carotid stenting, there was significant concern regarding increased risk of hemorrhagic complications.^10^ Second, TLs are considered a predictor of poor reperfusion after IVT alone due to underlying atherosclerosis that may impede successful reperfusion.^11,12^ Finally, in the setting of a TL caused by internal carotid artery (ICA) dissection, treatment with IVT raises concern about extension and worsening of the dissection.^13,14^

In acute LVO stroke, six randomized clinical trials (RCTs) have reported similar outcomes with IVT+MT versus MT alone.^15-20^ That said, the most recent guidelines from the European Stroke Organisation and Society of Vascular and Interventional Neurology still recommend that patients should receive IVT in addition to MT if eligible.^21,22^ In TLs, recent pooled analyses including the Thrombectomy In Tandem Lesions (TITAN) and Endovascular Treatment In Ischemic Stroke (ETIS) registries suggested that IVT prior to MT may increase the odds of favorable functional outcome without increasing the risk of hemorrhagic complications.^23^ Moreover, results from the German Stroke Registry-Endovascular Treatment study showed that the use of IVT in TL was an independent predictor of successful reperfusion (modified Thrombolysis in Cerebral Infarction [mTICI] score of 2b-3) in patients treated with MT.^6^ Nevertheless, the interaction between the regimen of intraprocedural APT and the risk of hemorrhagic complications after IVT was not assessed in these studies.

Hence, in this study, we sought to evaluate the safety and efficacy of MT with or without IVT for patients presenting with acute TLs. In addition, we assessed whether the use of intraprocedural APT was associated with a greater risk of clinically relevant intracranial hemorrhage when added to IVT.

## METHODS

### Study population

We used a pooled, international multicenter cohort registry for the study. Patient eligibility and methods of international collaboration have been reported previously.^24^ Briefly, the study included adult patients with TL treated with MT within 24 hours after stroke onset from 16 stroke centers (15 hospitals in the United States and 1 in Spain). TL was defined as an intracranial LVO (petrous, sigmoid, or terminus segment of the ICA or M1 or proximal M2 segment of the middle cerebral artery [MCA]) with concomitant extracranial ICA stenosis ≥50%.^25^ Patients were divided into two groups: 1) IVT + MT group (patients treated with IVT prior to MT), and 2) MT alone group (patients who did not receive IVT prior to MT). Treatment with IVT was determined at the discretion of the treating clinician. All intracranial occlusions were treated using a stent retriever and/or contact aspiration catheters. The endovascular and medical therapeutic interventions were performed according to the protocol of each institution under conscious sedation or general anesthesia and at the discretion of the neurointerventionalists. The study was approved under the waiver of informed consent by the local institutional review board at each participating center.

We classified the intraprocedural APTs into four categories depending on the intraprocedural APT regimen used during MT: no intraprocedural APTs, single oral APTs (aspirin, clopidogrel, or ticagrelor), dual oral APT (aspirin+clopidogrel or aspirin+ticagrelor), or intravenous APT (IV-APT) with or without oral APT (IIb/IIIa inhibitor alone; IIb/IIIa and single oral; IIb/IIIa and dual oral; cangrelor and single oral; cangrelor and dual oral).

### Outcome measures

The primary outcomes of the study were symptomatic intracranial hemorrhage (sICH) and parenchymal hematoma type 2 (PH2), as defined by the European Collaborative Acute Stroke Study (ECASS-3) criteria.^26^ We also assessed the rate of ischemic infarct hemorrhagic transformation defined as no hemorrhage, petechial hemorrhage (hemorrhagic infarction type 1 [H1] and type 2 [H2]), and parenchymal hematoma (parenchymal hematoma type 1 [PH1] and type 2 [PH2]), according to the Heidelberg Bleeding Classification.^27^

The secondary outcomes included successful (mTICI 2b-3) or complete reperfusion (mTICI 3), favorable functional outcome at discharge (discharge modified Rankin scale [mRS] 0-2), favorable functional outcome at 90 days (90-day mRS 0-2), excellent functional outcome at 90 days (90-day mRS 0-1), in-hospital mortality, and mortality at 90 days.

### Statistical analysis

Descriptive statistics were used to summarize continuous and categorical variables. We reported counts and percentages for categorical variables and means (SD) or medians (interquartile range [IQR]) for continuous variables. Shapiro-Wilk test and histograms were used to assess the normality of distributions. For the univariable analysis, we used the student’s t-tests or Wilcoxon rank-sum test for continuous variables and Chi-square or Fisher’s exact test for categorical variables, as needed.

To evaluate the safety and efficacy outcomes between the two patient groups, we performed multivariable logistic regression. Final models were selected by bidirectional stepwise selection procedures with Akaike’s information criteria or Bayesian information criteria, with candidate variables considered for inclusion in each model *a priori*: age, sex, hypertension, hyperlipidemia, atrial fibrillation, smoking status, previous stroke or transient ischemic attack, initial National Institutes of Health Stroke Scale (NIHSS), direct-to-angio suite strategy, mTICI 2b-3, intra-arterial tissue plasminogen activator, sICH, PH2, heparin, ICA treatment technique (stent-retriever vs non-stenting), ICA treatment timing (retrograde, anterograde, delayed), ethnicity, ICA stenosis/occlusion pre-procedure, etiology of ICA lesion, first pass effect, post procedure antiplatelets, early window (<6 hours from last known well [LKW]-to-arterial puncture) vs. late window (6-24 hours from LKW-to-arterial puncture), type of anesthesia, and Alberta Stroke Program Early Computed Tomography Score (ASPECTS). An adjusted multinomial regression model was also generated to estimate the odds of lower versus higher mRS scores at 90 days, and hemorrhagic transformation of infarction in an ordinal categorization, according to severity (none, petechial [H1, H2], parenchymal hematoma [PH1, PH2]). When fitting all the multivariable models, we included site as a random effect. The ICA treatment technique was forced into the models due to its association with functional outcomes and use of antiplatelet medication.^24^ Additionally, we explored effect modification by antiplatelet regimen for sICH, PH2, and hemorrhagic transformation. When an interaction was observed, we performed a sensitivity analysis for the subgroups.

We also performed sensitivity analysis by time window within 0-6 hours from LKW-to-arterial puncture (early window) for all the safety and efficacy outcomes and by type of admission on primary admission patients for favorable functional outcome. The 6 hours cutoff point was selected with the goal to include the larger proportion of eligible patients for IVT according to the current guidelines. ^28^

Lastly, we assessed the heterogeneity of the effect of IVT on pre-specified variables including procedural heparin, etiology, ASPECTS, intraprocedural antiplatelets, ICA treatment, time window, and ICA occlusion for the safety outcome (rate of sICH). We also looked at the effect of age, ASPECTS, intraprocedural antiplatelets, ICA treatment, time window, and ICA occlusion for the efficacy outcome mRS 0-2 at 90 days. The odds ratios (ORs) and 95% confidence intervals (CIs) for the effect size of each group were computed. All the statistical analyses were considered significant at a two-sided alpha level of ≤0.05. We used R statistical package (version 4.1.3, R Foundation for Statistical Computing, Vienna, Austria) for the analysis. Data will be made available upon reasonable request from the corresponding author.

## RESULTS

Of the 691 patients from the registry, 92 were excluded (**Figure 1**). Of the 599 patients included, 255 were in the IVT + MT group, and 344 were in the MT alone group. The demographic data and baseline characteristics of the two groups are presented in **Table 1**. Patients in the IVT + MT group were younger (66 vs. 69 years, *p* = 0.023), had a lower rate of hypertension (68.2% vs. 77.8%, *p* = 0.009), hyperlipidemia (41.7% vs. 50.1%, *p* = 0.042), atrial fibrillation (10.2% vs. 16.1%, *p* = 0.039), and history of antiplatelet medications (29.5% vs. 38.9%, *p* = 0.018) than those in the MT alone group. Furthermore, patients in the IVT + MT group had a higher median admission NIHSS (17 vs. 15.5, *p* = 0.027), a higher median ASPECTS (9 vs. 8, *p* < 0.01), a lower median number of MT passes (1 vs. 2, *p* = 0.01), higher rate of first-pass effect (66.9% vs. 55.8%, *p* = 0.007), a lower median time from LKW-to-reperfusion (300 vs. 657, *p* < 0.001), and a lower median time from LKW-to-arterial puncture (237 vs. 602 minutes, *p* < 0.001) than those in the MT alone group. Finally, 297 (50.4%) patients were treated within the early window, with a higher proportion of patients treated in the early window receiving IVT (81.1% vs. 27.2%, *p* < 0.001).

**Table 1.** Baseline and treatment characteristic of patients with endovascularly treated tandem lesions according to the use of intravenous thrombolysis.

|  | Total (N=599) | MT alone<br>(N=344) | IVT + MT<br>(N=255) | *p* value |
| --- | --- | --- | --- | --- |
| Age (years), median [IQR] | 68 [59–76] | 69 [61–76] | 66 [58–75] | **0.023** |
| Females, N° (%) | 191 (32) | 115 (33.4) | 76 (29.8) | 0.346 |
| White race, N° (%) | 375 (63) | 216 (63.5) | 159 (62.4) | 0.769 |
| Medical history, N° (%) |  |  |  |  |
| Hypertension | 440 (73.7) | 266 (77.8) | 174 (68.2) | **0.009** |
| Hyperlipidemia | 277 (46.6) | 171 (50.1) | 106 (41.7) | **0.042** |
| Diabetes | 168 (28.2) | 102 (29.8) | 66 (26) | 0.303 |
| Atrial fibrillation | 81 (13.6) | 55 (16.1) | 26 (10.2) | **0.039** |
| Current smoker | 141 (23.8) | 82 (24.3) | 59 (23.1) | 0.757 |
| Previous stroke/TIA | 93 (15.6) | 58 (17) | 35 (13.8) | 0.290 |
| Coronary artery disease | 109 (18.2) | 67 (19.5) | 42 (16.5) | 0.337 |
| Prior antiplatelets, N° (%) | 204 (34.9) | 130 (38.9) | 74 (29.5) | **0.018** |
| Transfer from outside institution, N° (%) | 327 (55.8) | 183 (54.8) | 144 (57.1) | 0.062 |
| Admission glucose (mg/dL), median [IQR] | 125 [107–154] | 125 [107–154] | 126.5 [107–154] | 0.610 |
| Baseline mRS, N° (%) |  |  |  | 0.306 |
| 0 | 466 (79.7) | 263 (78) | 203 (81.9) |  |
| 1 | 47 (8) | 26 (7.7) | 21 (8.5) |  |
| 2 | 41 (7) | 26 (7.7) | 21 (8.5) |  |
| 3 | 24 (4.1) | 18 (5.3) | 6 (2.4) |  |
| 4 | 5 (0.9) | 2 (0.6) | 3 (1.2) |  |
| 5 | 2 (0.3) | 2 (0.6) | 0 (0) |  |
| Admission NIHSS, median [IQR] | 16 [11–20] | 15.5 [10–20] | 17 [12–20.5] | **0.027** |
| ASPECTS, median [IQR] | 8 [7–9] | 8 [7–9] | 9 [7–10] | **<0.001** |
| Cervical ICA lesion etiology, N° (%) |  |  |  | 0.245 |
| Atherosclerosis | 470 (78.7) | 275 (80.4) | 195 (76.5) |  |
| Other | 127 (21.3) | 67 (19.6) | 60 (23.5) |  |
| Intracranial occlusion location, N° (%) |  |  |  | 0.111 |
| ICA | 93 (26.9) | 62 (30.4) | 31 (21.8) |  |
| M1 | 204 (59) | 118 (57.8) | 86 (60.6) |  |
| M2 | 49 (14.2) | 24 (11.8) | 25 (17.6) |  |
| Cervical ICA complete occlusion, N° (%) | 280 (51.9) | 167 (54) | 113 (48.9) | 0.238 |
| General anesthesia, N° (%) | 241 (40.2) | 141 (41) | 100 (39.2) | 0.662 |
| First-line technique, N° (%) |  |  |  | 0.468 |
| Combination | 310 (52.9) | 175 (52.1) | 135 (54) |  |
| Aspiration device | 177 (30.2) | 99 (29.5) | 78 (31.2) |  |
| Stent-retriever | 97 (16.6) | 60 (17.9) | 37 (14.8) |  |
| IA-tPA, N° (%) | 23 (3.9) | 14 (4.2) | 9 (3.6) | 0.723 |
| Number of passes, median [IQR] | 2 [1–3] | 2 [1–3] | 1 [1–3] | **0.001** |
| First pass effect, N° (%) | 351 (60.6) | 183 (55.8) | 168 (66.9) | **0.007** |
| ICA stenting, N° (%) | 365 (61.4) | 217 (63.5) | 148 (58.7) | 0.243 |
| Intraprocedural heparin, N° (%) | 218 (36.6) | 125 (36.5) | 93 (36.6) | 0.987 |
| Intraprocedural antiplatelets. N° (%) |  |  |  | 0.115 |
| Dual oral | 174 (29) | 102 (29.7) | 72 (28.8) |  |
| Any intravenous | 194 (32.5) | 119 (34.6) | 75 (29.4) |  |
| None | 106 (17.7) | 50 (14.5) | 56 (22) |  |
| Single oral | 125 (20.9) | 73 (21.2) | 52 (20.4) |  |
| Time metrics, median [IQR] |  |  |  |  |
| LKW-to-reperfusion (min) | 431 [274–769] | 657 [400–982] | 300 [236–399] | **<0.001** |
| LKW-to-arterial puncture (min) | 358 [211–706] | 602 [340–920] | 237 [169–334] | **<0.001** |
| Window, N° (%) |  |  |  | **<0.001** |
| Early | 297 (50.4) | 91 (27.2) | 206 (81.1) |  |
| Late | 261 (44.3) | 214 (63.9) | 47 (18.5) |  |
| Very late | 31 (5.3) | 30 (9) | 1 (0.4) |  |

IVT: Intravenous thrombolysis; MT: Mechanical thrombectomy; IQR: Interquartile range; TIA: Transitory ischemic attack; mRS: Modified Rankin scale; NIHSS: National Institutes of Health Stroke Scale; ASPECTS: Alberta Stroke Program Early Computed Tomography Score; ICA: Internal carotid artery; IA-tPA: Intraarterial tissue plasminogen activator; LKW: Last known well; min: minutes.

**Figure 1.**
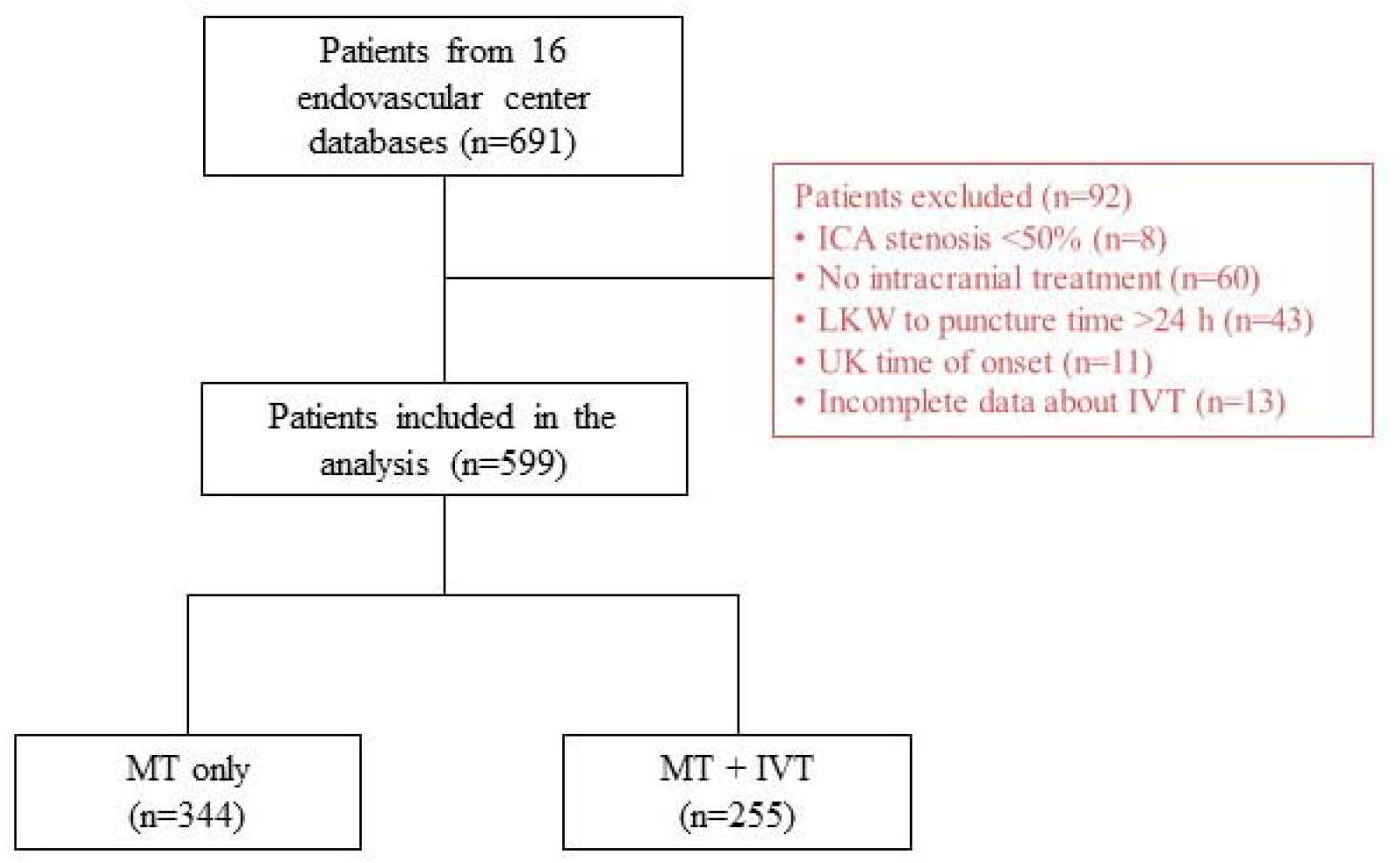
Flow diagram of patients included in the study. ICA: Internal carotid artery; LKW: last known well; UK: unknown; IVT: Intravenous thrombolysis; MT: mechanical thrombectomy.

### Primary outcomes

Compared with the MT alone group, the IVT + MT group had a higher rate of sICH, but this difference was not significant (7.5% vs. 5.3%, *p* = 0.273). After adjusting for the selected covariates, the difference in the risk of sICH between the two groups remained non-significant (aOR = 1.43; 95% CI 0.72–2.87; *p =* 0.308) (**Figure 2A**).

**Figure 2.**
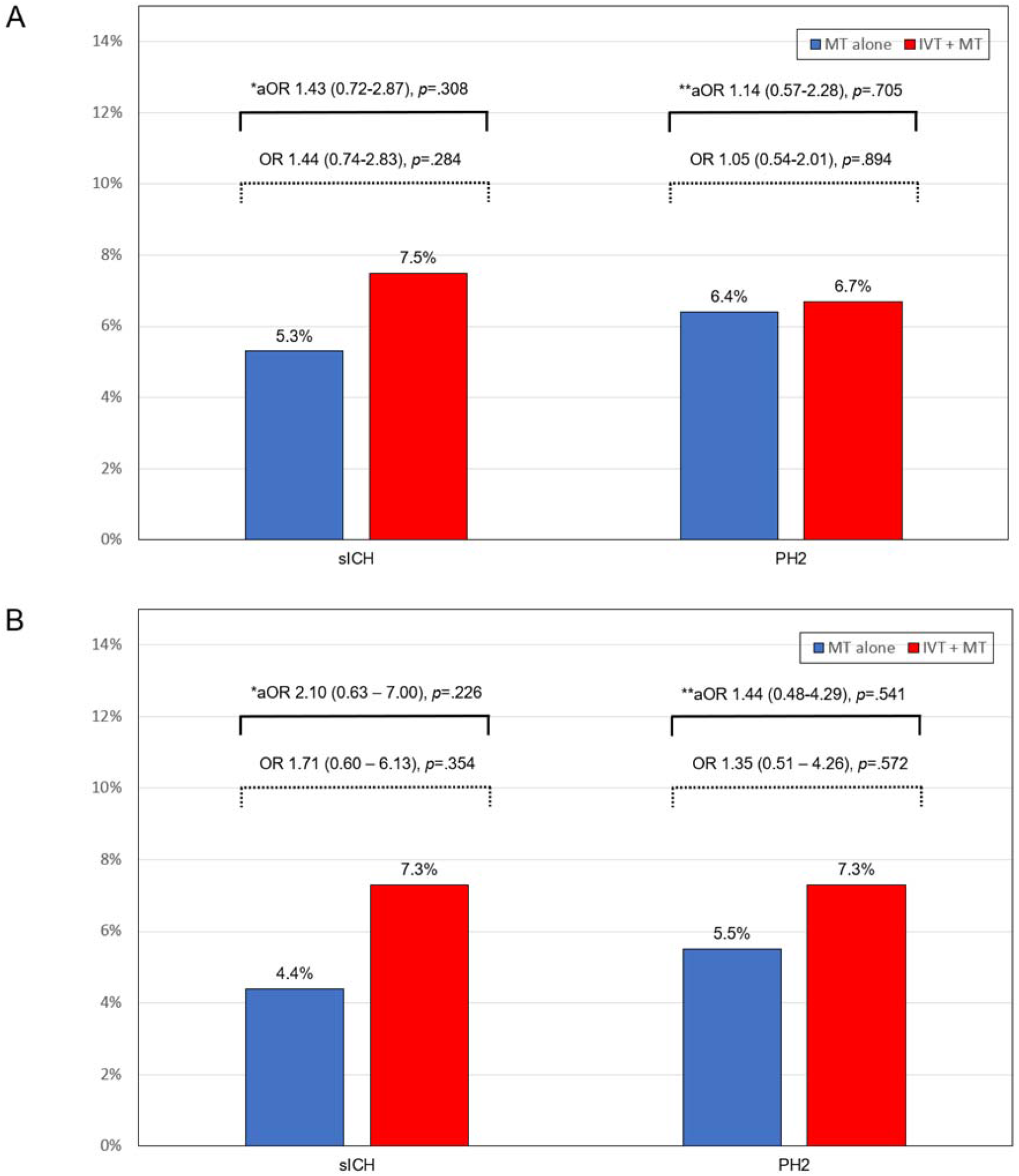
Bar chart of the (A) primary analysis and the (B) early window (0-6 hours) sensitivity analysis for symptomatic ICH and PH2 in patients treated with MT alone and IVT + MT. ICH: Intracranial hemorrhage; PH2: Parenchymal hematoma type 2; MT: Mechanical thrombectomy; IVT: Intravenous thrombolysis. *Adjusted for: ICA stenting, number of passes, mTICI 2b-3. **Adjusted for: ICA stenting, age, hypertension, ASPECTS.

When we assessed for any effect modification of intraprocedural APT on the relationship between IVT and sICH by including interaction terms in the model, a differential effect was observed between patients who received IV-APT and those who did not (*p* _interaction_ = 0.031). However, there was no evidence of heterogeneity in the remaining subgroups according to the use of IVT before MT (**Figure 3**). In the sensitivity analysis, IVT increased the risk of sICH in patients treated with IV-APT therapy (aOR = 3.58; 95% CI 1.17–10.89; *p =* 0.025). The effect of IV-APT on the increased risk of sICH with IVT appears related to the use of GP IIb/IIIa inhibitors (**Supplementary Figure 1**).

**Figure 3.**
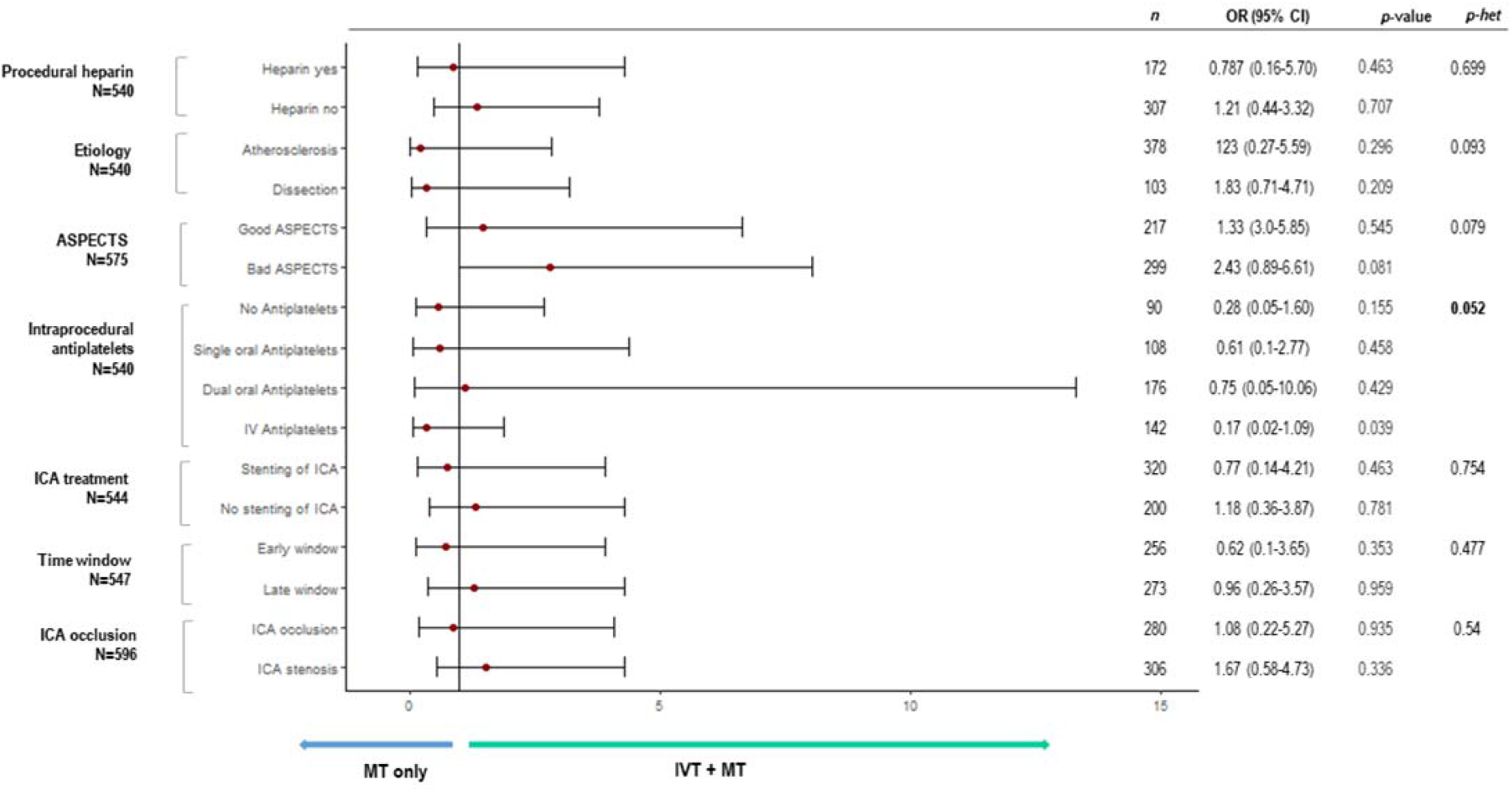
Comparisons in symptomatic intracranial hemorrhage according to the use of IVT in pre-specified subgroups. OR: Odds ratio for sICH; CI: Confidence interval; *p*-het: P value of heterogeneity; ASPECTS: Alberta Stroke Program Early CT Score; ICA: Internal carotid artery; MT: Mechanical thrombectomy. Adjusted for: number of passes, mTICI

There were no significant differences in the rates of PH2 between the two groups (6.7% vs. 6.4%, *p* = 0.894). After adjusting for confounders, the difference remained non-significant (aOR = 1.14; 95% CI 0.57–2.28; *p* = 0.705) (**Figure 2A**). The inclusion of interaction terms in the model to assess the effect of intraprocedural APT on the relationship between PH2 and IVT showed no differential effects (*p* _interaction_ > 0.05). In the sensitivity analysis, the use of IVT did not increase the risk of PH2 with any of the intraprocedural APTs.

With respect to hemorrhagic transformation, the risk was not significantly different between the MT alone and the IVT + MT groups (aOR = 0.92; 95% CI 0.54–1.57; *p* = 0.751) (**Supplementary Figure 2**). The interaction evaluation showed no significant effect between intraprocedural APT and IVT for hemorrhagic transformation (*p* _interaction_ > 0.05). In the sensitivity analyses, the use of IVT did not increase the risk of hemorrhagic transformation with any of the intraprocedural APTs.

### Secondary outcomes

At 90 days, there was a higher, albeit not statistically significant, rate of favorable functional outcome in the IVT + MT group compared with the MT alone group (49.2% vs. 45.2%, *p* = 0.363). After adjusting for covariates, the IVT + MT group had higher odds of a favorable functional outcome (aOR = 1.76; 95% CI 1.05–2.94; *p =* 0.030) (**Figure 4A**). In addition, we observed a trend toward increased odds of a favorable outcome in primary admission patients treated with IVT (aOR = 2.12; 95% CI 1.00 – 4.64; *p* = 0.054). There was no evidence of heterogeneity in subgroup differences according to the use of IVT before MT (**Figure 5**).

**Figure 4.**
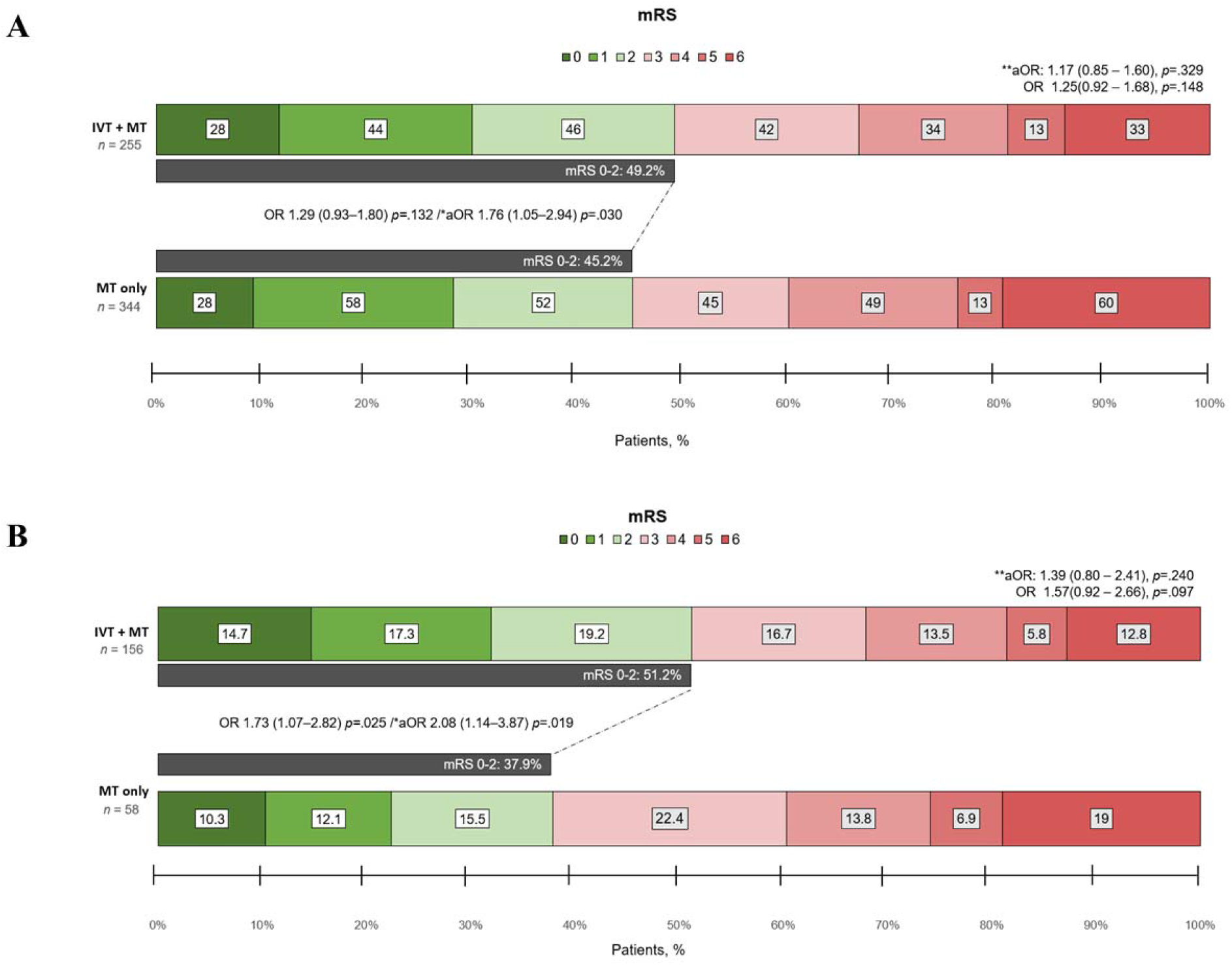
A) Bar chart of shift analysis of modified Rankin Scale (mRS) at 90 days of the entire cohort. B) Sensitivity analysis for patients within 6-hour time window only. aOR: Adjusted odds ratio; CI: Confidence interval; IVT: Intravenous thrombolysis; MT: Mechanical thrombectomy. *Categorized mRS 0-2 vs. 3-6. Adjusted for age, NIHSS, type of anesthesia, successful reperfusion, internal carotid artery stenting, symptomatic intracranial hemorrhage, and postprocedural antiplatelet therapy. **Multinomial model. Adjusted for age, NIHSS, type of anesthesia, successful reperfusion, internal carotid artery stenting, symptomatic intracranial hemorrhage, and postprocedural antiplatelet therapy.

**Figure 5.**
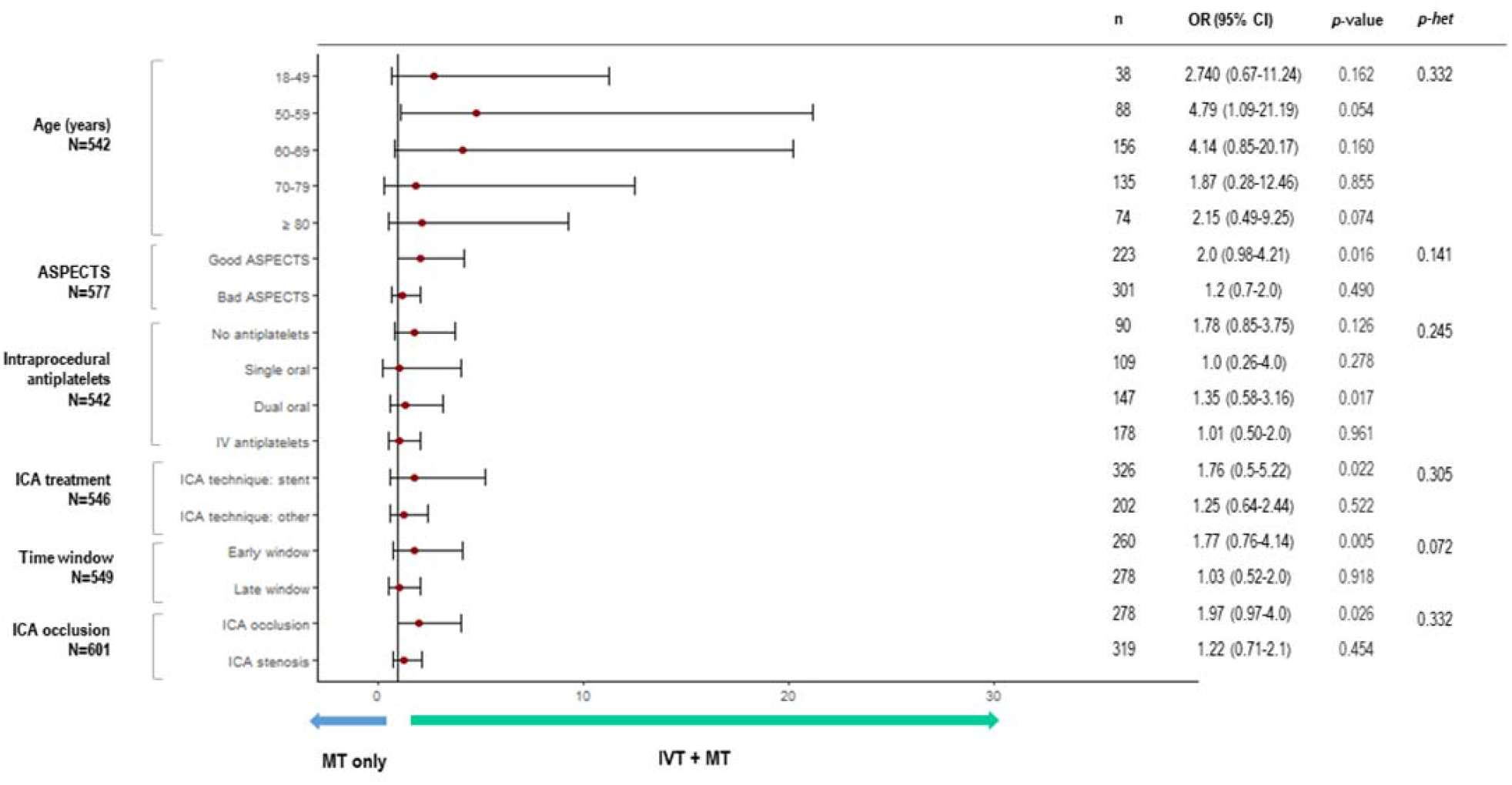
Comparisons of favorable outcomes at 90 days according to the use of IVT in pre-specified subgroups. OR: Odds ratio of favorable outcome at 90 days; CI: Confidence interval; *p*-het: P value of heterogeneity; ASPECTS: Alberta Stroke Program Early CT Score; ICA: Internal carotid artery; MT: Mechanical thrombectomy.

The odds of successful reperfusion (aOR = 1.03; 95% CI 0.53–2.00; *p* = 0.928) and excellent reperfusion (aOR = 0.93; 95% CI 0.63–1.38; *p* = 0.729) were not different between patients treated with IVT+MT versus MT alone (**Table 2**). There was no significant interaction between intraprocedural APT and IVT for successful reperfusion (*p* _interaction_ = 0.112); however, there was a significant interaction between IV-APT use with IVT for the outcome of excellent reperfusion (*p* _interaction_ = 0.034). In the sensitivity analysis, the use of IVT did not increase the odds of achieving excellent reperfusion with any of the intraprocedural APTs.

**Table 2.**
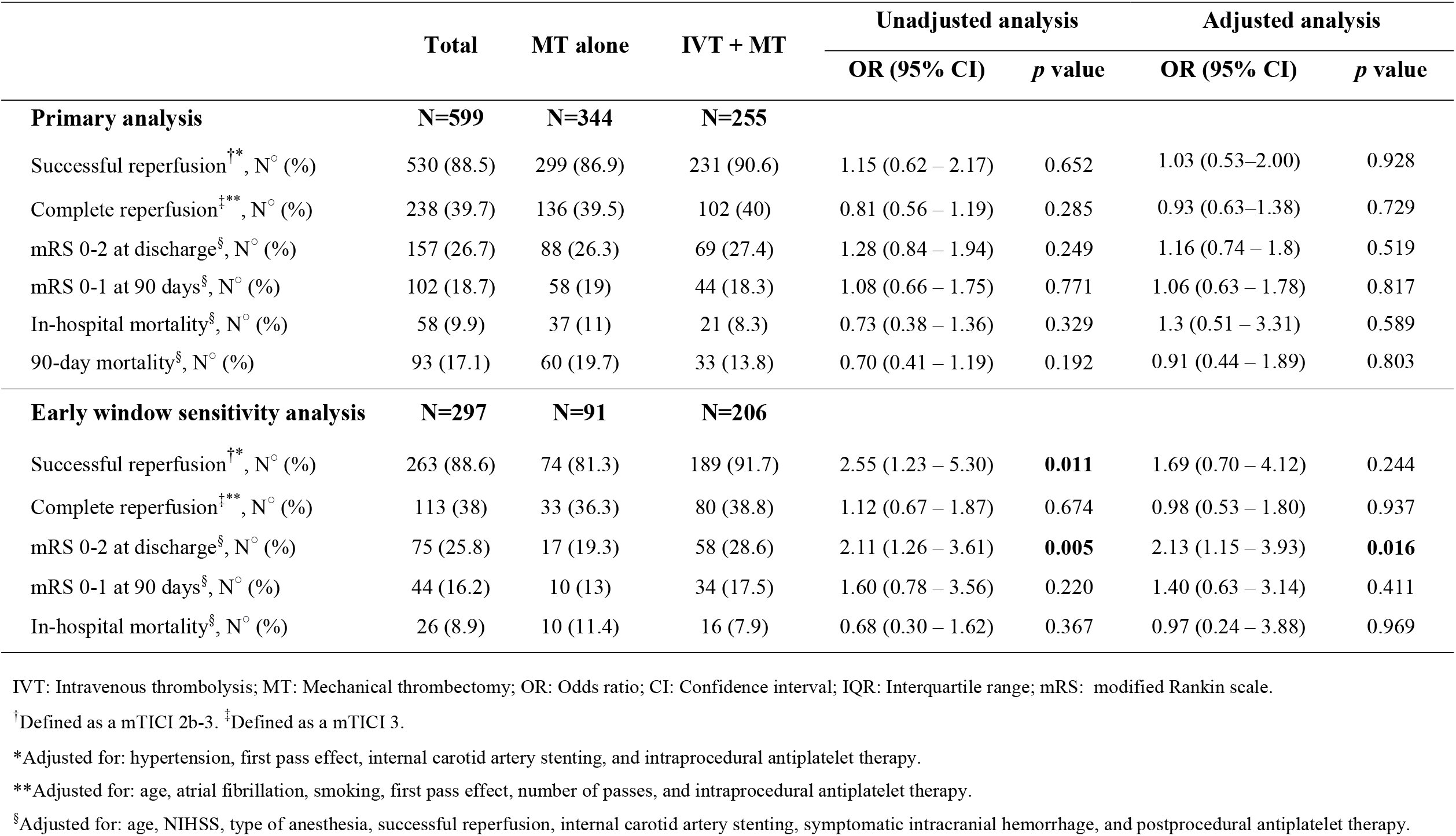
Secondary outcomes of patients with endovascularly treated tandem occlusion stroke according to the use of intravenous thrombolysis.

In the multivariable analysis, no association was found for in-hospital mortality (aOR = 1.30; 95% CI 0.51 – 3.31; *p* = 0.589) and 90-day mortality (aOR = 0.91; 95% CI 0.44 – 1.89; *p* = 0.803) with the use of IVT prior MT (**Table 2**).

### Early window sensitivity analysis

Early window included only patients with LKW-to-arterial puncture time < 6 hours. Similar to the primary analysis, we found no significant differences on the rates of sICH (aOR = 2.18; 95% CI 0.68 – 7.05; *p* = 0.192), PH2 (aOR = 1.44; 95% CI 0.48 – 4.29; *p* = 0.514) (**Figure 2B**), and hemorrhagic transformation (aOR = 1.25; 95% CI 0.71 – 2.19; *p* = 0.444). On the other hand, the IVT + MT group showed greater odds of a favorable functional outcome at discharge (aOR = 2.13; 95% CI 1.15 – 3.93; *p* = 0.016) and at 90-days (aOR = 2.08; 95% CI 1.14 – 3.87; *p* = 0.019) (**Figure 4B**). Finally, there were no differences for additional outcomes (**Table 2**).

## DISCUSSION

Our study compared the safety and efficacy between IVT+ MT versus MT alone in patients presenting with acute TLs. We found that (1) the administration of IVT before MT was safe and was not associated with an increased risk of sICH, PH2, or hemorrhagic transformation; (2) treatment with IVT plus MT was associated with a higher rate of favorable functional outcome at 90 days, especially in TL-patients treated within the 6-hour time window; and (3) the use of intraprocedural IV-APT was associated with an increased the risk of sICH in patients who received IVT.

The role of IVT in LVO stroke patients eligible for MT has been a subject of debate. Theoretically, adding IVT may contribute to achieving early reperfusion of the ischemic territory before MT^12,29-31^, increase reperfusion rates with fewer recanalization attempts^32^, and may improve outcomes in patients with failed MT reperfusion attempts^33^. However, the theoretical risk of distal clot embolization to locations not amenable to MT ^34^ and intracranial hemorrhage, the potential delays for arterial puncture, and its elevated cost are considerable disadvantages.^35,36^ Six recent RCTs evaluating the use of IVT+MT procedures for LVO have shown similar rates of 90-day functional outcomes and sICH when compared to MT alone.^15-20^ Consequently, evidence-based recommendations by the European Stroke Organization (ESO)-European Society for Minimally Invasive Neurological Therapy (ESMINT), and Vascular + Interventional Neurology (SVIN) recommend the use of IVT for eligible patients.^21,22^

TLs are often excluded and, therefore, underrepresented in clinical trials. As a result, the level of evidence to evaluate the safety of the use of IVT+MT is limited. A pooled analysis of the TITAN and ETIS registries found that bridging therapy did not increase the risk of sICH or PH2, as per our findings.^23^ They reported a rate of 7.5% for sICH and 5.6% for PH2 in their IVT+MT group which is comparable with our 7.5% rate for sICH and 6.7% for PH2. Furthermore, our rate of sICH was similar to the rates reported in the six RCTs of bridging therapy in acute LVO, which ranged from 4.7% to 7.8%.^15-20^ In our study, we also found that the IVT+MT did not increase the risk of hemorrhagic transformation, which is relevant considering that IVT may promote hemorrhagic transformation through fibrinolytic and/or immune mechanisms.^37^ This finding was consistent in our population independently of the underlying etiology, baseline ASPECTS, use of stenting, complete cervical occlusion or presentation time.

APT has been reported safe, with a low risk for intracranial hemorrhage in patients with TL.^10^ Its administration primarily occurs intraprocedural to prevent an acute in-stent thrombosis and/or subsequent restenosis of the cervical segment when a cervical ICA stenting is pursued.^38^ According to the ATRIS trial results, early therapy with intravenous aspirin may increase the risk of hemorrhagic complications in patients who have already received IVT.^39^ On the contrary, Zhu *et al*. found that IVT prior to MT did not lead to a significant association with APT (aspirin, clopidogrel, or glycoprotein IIb/IIIa receptor antagonist) and hemorrhagic and/or procedural complications in patients with TL.^9^ Similarly, Anadani *et al*. did not find evidence of heterogeneity in treatment effect sizes according to prior IVT for the risk of hemorrhagic complications. They concluded that ICA stenting was safe in patients with previous IVT.^4^ In our study, we found an IVT-by-intraprocedural APT interaction. Intraprocedural IV-APT (which included GP IIb/IIIa inhibitors and cangrelor) seemed to increase the risk of sICH in patients treated with IVT+MT. Similarly, Stampfl *et al*. attributed the high ICH rate (16.6%) to intravenous APT with tirofiban, considering that most of their patients (92%) were treated with IVT prior to ICA stenting.^40^ Also, Heck and Brown found that intravenous APT with abciximab after acute ICA stenting may be associated with higher rates of sICH (31%) in TLs,^41^ and one matched cohort analysis reported a higher rate of parenchymal hemorrhage in patients with TLs and those concomitantly treated with eptifibatide.^42^ The ATILA project (https://www.clinicaltrials.gov; unique identifier: NCT05225961) is a multicenter phase IV RCT aimed at determining the safety and efficacy of intravenous tirofiban versus intravenous aspirin in patients with TL treated with MT. The results of this trial will be helpful in understanding the safety of tirofiban in TL. In addition, we observed that the use of GP IIb/IIIa inhibitors was associated with all the hemorrhage cases related to IVT compared with cangrelor (**Supplementary Figure 1**). However, the small sample sizes in these subgroups are a limitation for drawing a definite conclusion.

We did not find an association between the use of IVT and successful reperfusion when evaluating our entire TL population. These results are in contrast with the recently published literature that evaluated TL presenting within 8 hours after symptom onset.^23^ However, when evaluating patients in the early window after adjusting for confounders, IVT was associated with higher odds of functional independence at 90 days. Interestingly, the different effects of IVT on successful reperfusion and functional independence reflect the heterogeneity and chronicity of the occlusions, which may introduce further complexity in the treatment of TLs due to collateral circulation. More importantly, the advantages of pretreatment with IVT, such as clot decomposition and recanalization of microvasculature, might have an additional beneficial effect independently of proximal vessel recanalization.^43^ In fact, the recently reported CHOICE trial has shown that the addition of intra-arterial tissue plasminogen activator was associated with better functional outcomes at 90 days without any significant differences in the rates of reperfusion or sICH.^44^

This study has several limitations. First, there is a potential selection bias due to the retrospective nature of its design. Second, the patient selection was determined according to the clinical evaluation of each center and neuro-interventionalist and, therefore, lacks randomization. Third, the clinical and imaging data were self-adjudicated by independent investigators at each center without external control or core imaging laboratory adjudication. Finally, the predictive models should be interpreted with caution considering the limited sample size and the potential risk of confounding by measured and unmeasured variables.

## Data Availability

Data not provided in the article because of space limitations may be shared (anonymized) at the request of any qualified investigator for purposes of replicating procedures and results.

## CONCLUSIONS

Our study shows that IVT+MT for TL cases did not increase the overall risk of sICH, PH2, or overall hemorrhagic transformation independently of the cervical revascularization technique used. However, the use of intraprocedural IV-APT during stent implantation may be associated with an increased risk of sICH in patients who received IVT prior to MT. Importantly, treatment with IVT+MT was associated with a higher rate of favorable functional outcomes at 90 days, especially in patients within the 6-hour window. Prospective studies are warranted for confirmation.

## Acknowledgements

We would like to acknowledge the followings for their contribution: Kathie Gonzales, CRA, University of Iowa Hospitals and Clinics.

